# Mpox knowledge, behaviours and barriers to public health measures among gay, bisexual and other men who have sex with men in the UK: A qualitative study to inform public health guidance and messaging

**DOI:** 10.1101/2023.05.19.23290102

**Authors:** Tom May, Lauren Towler, Louise E Smith, Jeremy Horwood, Sarah Denford, G James Rubin, Matthew Hickman, Richard Amlôt, Isabel Oliver, Lucy Yardley

## Abstract

**Background:** The 2022-23 Mpox epidemic is the first-time sustained community transmission had been reported in countries without epidemiological links to endemic areas. During that period, the outbreak almost exclusively affected sexual networks of gay, bisexual, or other men who have sex with men (GBMSM) and people living with HIV. In efforts to control transmission, multiple public health measures were implemented, including vaccination, contact tracing and isolation. This study examines knowledge, attitudes, and perceptions of Mpox among a sample of GBMSM during the 2022-23 outbreak in the UK, including facilitators for and barriers to the uptake of public health measures.

**Methods:** Interviews were conducted with 44 GBMSM between May and December 2022. Data were analysed using reflexive thematic analysis. Positive and negative comments pertaining to public health measures were collated in a modified version of a ‘table of changes’ to inform optimisations to public health messages and guidance.

**Results:** Most interviewees were well informed about Mpox transmission mechanisms and were either willing to or currently adhering to public health measures, despite low perceptions of Mpox severity. Measures that aligned with existing sexual health practices and norms were considered most acceptable. Connections to GBMSM networks and social media channels were found to increase exposure to sexual health information and norms influencing protective behaviours. Those excluded or marginalized from these networks found some measures challenging to perform or adhere to. Although social media was a key mode of information sharing, there were preferences for timely information from official sources to dispel exaggerated or misleading information.

**Conclusions:** There are differential needs, preferences, and experiences of GBMSM that limit the acceptability of some mitigation and prevention measures. Future public health interventions and campaigns should be co-designed in consultation with key groups and communities to ensure greater acceptability and credibility in different contexts and communities.

## Introduction

Mpox (formerly human monkeypox) is a zoonotic orthopoxvirus with clinical features similar to smallpox (e.g., fever, lesions), although with substantially lower mortality (1, 2). Since human cases were first detected in 1970, sporadic outbreaks and cases have been reported in regions of West and Central Africa where the disease is endemic (3–5). Traditionally, transmission has occurred via contact with infected animal reservoirs, although evidence of human-to-human transmission has been reported in recent outbreaks in Nigeria (4) and the Democratic Republic of Congo (5, 6). Prior to 2022, cases outside of endemic areas were rare and linked to contact with imported animals (7) or travel to endemic areas (8–11).

In May 2022, cases of Mpox were detected in the UK, with clusters soon reported in multiple non-endemic countries. As of 27^th^ April 2023, 87,113 cases have been reported in 111 countries, including many without previously documented cases of Mpox (12). This is the first-time sustained community transmission has been reported in countries without epidemiological links to endemic areas. The outbreak was declared a public health emergency of international concern by the World Health Organization (WHO) on July 23^rd^, 2022. Since the global peak of 7576 cases observed in the week of 8^th^ August 2022, the number of cases reported has declined substantially: between 30^th^ January 2023 and 23^rd^ April 2023, the average number of global cases observed weekly was 136 (12).

Internationally, the 2022-23 Mpox outbreak has almost exclusively affected sexual networks of gay, bisexual or other men who have sex with men (GBMSM) (84.1%) and people living with HIV (PLHIV) (48.5%), with prolonged close or intimate contact with an infected individual the primary route through which infection occurs (12, 13). There are some reports of clusters associated with sex-on-premises venues or sex parties, which has prompted debates as to whether Mpox should be considered a sexually transmitted infection (STI) (14). Novel clinical characteristics and transmission – including anogenital and oral mucosal lesions presenting at inoculation sites and Mpox DNA detected in seminal fluid (15) – support the potential role of sexual contact as a driver of transmission in the outbreak (13).

The UK has the eighth-highest case rate of mpox of all nations and the third-highest in the European WHO region (12). A national response, composed of a range of public health interventions, was enacted by the United Kingdom Health Security Agency (UKHSA) in efforts to control transmission. This included diagnostic testing for suspected cases, 21 days self-isolation for confirmed cases, and tracing contacts of confirmed cases. Contacts were managed according to exposure risk with smallpox (modified vaccinia Ankara, MVA) vaccination recommended for those at highest risk of Mpox exposure, including GBMSM who attend sex on-premises or engage in group sex or sex with multiple sexual partners. People were also advised to contact a sexual health clinic if they had suspected Mpox symptoms and had either been in close contact with a confirmed or suspected Mpox case, or if they had recently travelled to central or West Africa (16).

The public response to these measures, including their acceptability among GBMSM, has been evidenced. In the UK, a cross-sectional survey (n=1932) of GBMSM and the general population found high levels of vaccine acceptability (86%) and fairly high self-reported intention to self-isolate (61%) (17). Understandings of public health information (including Mpox symptoms, origins and where to attend if symptomatic) were more varied, with the most trusted sources of information found to be healthcare professionals (37%), official health agencies (29%) and mainstream media (12%). A similar study conducted with GBMSM and the general population in the UK also found intentions to enact protective behaviours (including help seeking, reducing sexual contact, contact sharing, isolation and vaccination) to be high, with GBMSM more likely to report intending to enact behaviours (except for self-isolation) (18). Greater intention to engage with behaviours and targeted public health measures were associated with perceived susceptibility to and severity of Mpox among general population and GBMSM samples. Similar patterns of vaccine acceptability have been observed in GBMSM in the WHO European Region, with willingness to accept a vaccine associated with perceived severity of and susceptibility to Mpox (19), and being single but dating or in an open relationship (20). Being linked to routine sexual healthcare through recent STI diagnosis or PrEP/antiretroviral use was also associated with increased willingness to receive a vaccine (19).

Whilst the acceptability of measures is likely to have contributed to both recent decreases in STIs and Mpox transmission in England among GBMSM (18), some may still find measures difficult to follow. For example, those from the lowest income backgrounds and minoritized populations were found to face additional barriers to adherence to mitigation measures during the COVID-19 pandemic (21). Vaccine acceptability and uptake were also lower among these groups (22–25), with mistrust, concerns about side effects and accessibility factors (e.g. language barriers) identified as common reasons (22, 24, 26). There is evidence that similar patterns affected the uptake of measures to prevent the spread of Mpox, with minoritized GBMSM groups and those unable to afford basic needs being less inclined to receive a vaccine (17). Evidence from the United States suggests vaccine uptake among GBMSM has been differential, with Black and Hispanic people less likely to receive a vaccine despite accounting for a large proportion of cases (27). Hence, there are concerns that Mpox may further expose and amplify existing health disparities and inequalities (28).

Despite an emerging body of quantitative work, there is no published qualitative research into the 2022-23 Mpox outbreak. Qualitative insights into acceptability and feasibility of public health advice for all target users are particularly important to understand potential reasons for non-adherence to advice. Hence, this study qualitatively describes knowledge, attitudes and perceptions of a sample of GBMSM toward Mpox during the 2022-23 outbreak in the UK, including facilitators for and barriers to the uptake of transmission-reducing behaviours (e.g., healthcare seeking/testing, self-isolation, vaccination). This study also explores preferences for optimising and adapting public health messaging to improve the uptake of measures.

## Methods

### Design

The research employed a qualitative design using semi-structured remote telephone/video-call methods with 44 GBMSM. Interviews were conducted between June and December 2022. The study used elements of the Agile Co-production and Evaluation (ACE) Framework, a novel approach to rapidly developing public health interventions, messaging and guidance. Specific ACE methods are published elsewhere (29), but elements used in this work are outlined below.

### Sample and recruitment

Eligibility was based on evolving needs and initially comprised confirmed cases of Mpox, and people identified at higher risk of Mpox, including healthcare workers. We therefore conducted a small number of interviews with cis-women healthcare workers (n=2) during the initial outbreak period (not included in this paper). However, as the epidemic unfolded it became apparent that cases of Mpox were primarily focused occurring among GBMSM. All subsequent interviews were therefore conducted with GBMSM, including some cis-gendered men identifying as heterosexual but reporting having sex with men. Being over 18 years of age and currently living in the UK were also eligibility requirements.

In line with the ACE framework, the study used multiple online and offline recruitment methods to ensure representation from a diverse sample in terms of ethnicity, educational level, Mpox status, geographic location, and social and sexual practices. This included the dissemination of recruitment material on both personal and organizational social media accounts (Facebook, Twitter and Instagram) and a location-based online dating application (Grindr) targeted at postcodes in Manchester and Brighton to ensure geographical variation. The research team also utilized existing partnerships and contacts with sexual health and LGBTQ+ community-based organizations, who shared study material with eligible interviewees through their regular online and face-to-face outreach work, including at Pride events in Summer 2022. These organizations also reached out to specific groups under-represented in our research, including those from minoritized ethnic groups or whose first language was not English, as well as those at increased risk of Mpox through attendance at saunas or sex-on-premises venues.

Individuals registered their interest via an online sign-up page (hosted by Qualtrics) and were asked to leave contact and demographic details. Recruitment for the study commenced in May 2022, shortly after the first reported Mpox case in the UK on 6^th^ May 2022.

Measures were also undertaken to screen out several participants identified as likely fraudulent. For example, prior to interviews the research team were made aware of several ‘red flags’ used by other qualitative researchers to identify fraudulent research participation, including participants not turning on cameras, brief and vague responses to questions, the citation of recruitment sites not used in the study and frequent emails post-interview requesting payment (30, 31). Similar instances occurred in this study among several interviewees identifying as heterosexual (we did not explicitly screen sexuality prior to interview) reporting recent Mpox acquisition. Vague responses were also given in response to questions regarding acquisition (including no mention of sexual contact with confirmed cases or travel to endemic areas), diagnosis, symptoms and knowledge of Mpox measures for confirmed cases (e.g., isolation periods). Given how the 2022-23 Mpox outbreak almost exclusively affected sexually active networks of GBMSM and the increasing incidence of ‘imposter’ or deceitful participants motivated by financial incentives to participate in qualitative studies (30), we became suspicious about the authenticity of these interviewees. We therefore chose to exclude these interviewees from the study following interview to maintain the integrity of the research.

Ethical approval was granted by the UK Health Security Agency Research Ethics and Governance Group: Reference R&D 512.

### Data collection

Interviews were conducted by TM (research fellow in behavioural science) and LT (lecturer in health psychology) via telephone or video call. Both authors have experience in sexual health and sex research. A flexible topic guide (see supplementary material) with open-ended questions was used to elicit views of Mpox (including perceptions of risk and severity) and perceptions of and reactions to mitigation measures aimed at reducing transmission, including isolation, contact tracing, vaccination and healthcare seeking behaviours. Following this, interviewees were shown official UKHSA posters (see supplementary material) and asked what they thought of the content. Think aloud questions (e.g., “what do you think of this message?”) were used to prompt suggestions for the optimization of messages and content. All interviewees were offered £20 in cash or vouchers for each 30 minutes of interview time.

### Data Analysis

With consent, interviews were recorded, transcribed, anonymized, and uploaded to NVivo V.12 software for analysis through a reflexive thematic approach taking a critical realist perspective (32, 33). This began with TM and LT independently reading and coding the same three transcripts. A preliminary coding framework, informed deductively by concepts within the topic guide and components from Protection Motivation Theory (PMT) (e.g. perceived susceptibility and severity of Mpox and perceived effectiveness and self-efficacy for protection measures, e.g. testing, self-isolation, contact tracing and vaccination) (34) were used to guide the coding of these transcripts. An inductive approach was then used to refine the framework to reflect any themes or concepts within the data and applied to the remaining transcripts by LT, who coded and synthesised text into categories, which were subsequently analysed and grouped into themes. To ensure rigour in this process, TM and LT met weekly to discuss and iteratively refine new codes or themes that were of potential significance to the research objectives.

### Optimisations to Public Health Messaging

To identify any suggested optimisations to official UKHSA posters, we collated all positive and negative comments pertaining to materials in a modified version of a ‘table of changes’ (TOC). The TOC is a tool commonly used in the person-based approach (PBA) (35) whereby quotes relating to elements of an intervention content (including messaging) are used to identify possible intervention-specific optimizations required to promote engagement and behaviour change. To maximise the effectiveness of this approach in a rapid optimisation context (see also (36)), we used a hybrid method that integrated insights from the thematic analysis into the TOC. Hence, perceptions regarding risk and severity of Mpox and perceived acceptability of and capacity to perform public health measures were linked to intervention content in the TOC. For example, quotes relating to one’s perceived risk of contracting Mpox were connected to messaging content around risk reduction, whilst barriers influencing engagement with specific measures were linked to optimisations concerning guidance about specific protective behaviours. This allowed general information relating to Mpox knowledge, attitudes, and behaviours to be collated alongside specific feedback on posters and messages obtained in the interviews, thereby ensuring broader social determinants and contexts influencing behaviour were incorporated into and accounted for in the intervention optimisation processes.

## Results

44 interviews were conducted with GBMSM with an average age of 34.2 years (range 20-56). The majority of interviewees were White British (n = 21), followed by White Other (n=8). 13 interviewees reported recent contact with a confirmed Mpox case, and eight reported recent Mpox diagnosis.

The thematic analysis identified two main themes: (1) *Perceived Risk and Severity of Mpox* and (2) *Perceived Acceptability of and Capacity to Perform Measures*. Feedback on UKHSA public health measures were also obtained and helped inform optimisations to overcome any behavioural barriers identified in the thematic analysis. These are presented in the third theme, *Optimisation of Messaging*, which includes the TOC.

### Perceived Risk and Severity of Mpox

Interviewees’ assessments of their level of risk played an important role in willingness to engage with some measures, including healthcare seeking and vaccination uptake. As sexual contact was widely recognized as the dominant mode of transmission, perceived risk was strongly influenced by recent sexual practices or activity. Those who attended sex-on-premises venues or reported recent casual sex recognized they were at greater risk of acquisition, and were generally highly motivated to engage with measures to reduce the level of risk to both themselves and others. This included avoiding sexual contact until they received a vaccine (*Until I had my vaccination I avoided sexual contact with anybody and everybody. I basically just switched it off; I was like, that is just too risky,* Aged 46-50, White British) and seeking healthcare if they had recent sexual contact with a confirmed case:

> *So I had loads of sex in the dark room with some total strangers…then on Monday I got a text from one of the people that I had met that night saying that he had a fever on the Sunday…I realised that it would make sense to call myself in [to STI clinic]* (Aged 26-30, White British)

The possibility of severe health complications also influenced some interviewees’ perceptions of risk and subsequent decisions to enact risk-reducing behaviours. This was most pronounced among those with comorbidities concerned about the possibility of severe Mpox infection or the exacerbation of their condition. One PLHIV described how this influenced his decision to enact ‘careful’ practices to prevent future health complications:

> *I am diabetic and have HIV so being somebody that lives with two conditions that can be complicated by other things, I suppose I’m more hyperaware and nervous about it [Mpox] because of that…I am thinking about it when meeting people and putting myself at risk. I’ve definitely been more careful with meeting people* (Aged 46-50, Mixed Other)

In contrast, although non-immunocompromised interviewees expressed concern about scarring from lesions and severe pain, Mpox was not considered a significant health concern. The availability of a vaccine and a perception that Mpox was non-life threatening and curable were the most common reasons given. Most interviewees also considered themselves to be relatively young and would not become seriously ill if infected:

> *I’m not really concerned about getting Monkeypox because I seem to understand it’s not something that’s dangerous, it’s just something that you don’t really want…personally it’s not a massive concern. I would not want it but if I were to catch it, I don’t think it could be the end of the world* (Aged 31-35, White Other)

Low perceived Mpox severity could also be understood within a context of ongoing assessment and appraisal of other health threats. There was a perception that, in relation to more ‘severe’ STIs, including HIV, Mpox was less serious and could be quickly identified and treated through frequent STI testing. Some PrEP users described how the benefits of protection from HIV therefore undermined protection from ‘less severe’ STIs, including Mpox:

> *I related a lot to HIV risk decisions because I’m on PrEP, how I started feeling a lot more comfortable with deciding not to use condoms, not to wear condoms after taking PrEP and even though I’m very much aware that there’s still a lot of risks with other STIs, it always seems that it’s still worth the risk. Just because of the availability of treatment and it’s very easy to get tested, and symptoms are not severe* (Aged 31-35, Latin American)

Perceived Acceptability of and Capacity to Perform Measures

Specific barriers relating to each mitigation measure were identified and can be found in Table 2. Three main issues were identified across all measures and are discussed below.

**Table 1.** Demographic characteristics of interviewees

| **Demographics** | **Range/n (mean)** |
| --- | --- |
| **Age (years)** | 20-56 (34.2) |
| **Ethnicity** |  |
| White British | 21 |
| White Other* | 8 |
| Black / Black African / Black Caribbean | 5 |
| Latin American, inc mixed | 5 |
| Asian | 3 |
| Other | 2 |
| **Gender** |  |
| Cis male | 43 |
| Gender queer | 1 |
| **Sexual Orientation** |  |
| Gay/homosexual | 40 |
| Other, inc queer | 4 |
| **Highest Educational Qualification** |  |
| Postgraduate studies completed | 20 |
| Undergraduate studies completed | 14 |
| Currently in full time education | 6 |
| 6<sup>th</sup> form/college/equivalent completed | 4 |
| **Recent contact with confirmed Mpox case** |  |
| No | 24 |
| Yes | 13 |
| Don't know | 7 |
| **Confirmed Mpox case** |  |
| No | 36 |
| Yes | 8 |

\*Includes White European

**Table 2.** Key barriers potentially influencing engagement with specific measures

| Mitigation measures | Barrier Identified | Quotes |
| --- | --- | --- |
| Symptom recognition, healthcare seeking and testing | Stigma | What would I do in that situation? I would be very embarrassed, very, very ashamed. I think I really had my own little scare, a couple of weeks back and I was like, 'Have I?' I didn't get tested. I didn't do it because I just felt so – I was riddled with shame about it (Aged 26-30, White Other). |
|  | Consequence of reactive test (e.g., prolonged isolation period) | Someone might be like, okay I'm not feeling good at the moment but I've got Glastonbury in two weeks and a half or I'm going on holiday...so if I say that now it means that I can't go to this plan that I've had for ages and I've been looking forward to so I have to take it into my own hands (Aged 31-35, White British). |
|  | Lack of specific symptoms | I guess it's tricky when the symptoms are so generic... generic symptoms like that I don't think I would necessarily get in touch with anyone (Aged 21-25, White British) |
| Contact tracing | Not knowing contact information | The people on the phone, it was so strange in that they seemed to think that you would have all the details of everyone you had sex with such as full name, date of birth, address and NHS number (Aged 26-30, White British) |
|  | Stigma and awkwardness | I think it's always an awkward one because obviously so many of these people are probably going to be sexual contacts as well, so it's about putting that stuff out there and making that public to other people which is always an awkward one (Aged 46-50, Mixed Other) |
| Isolation | Impact on wellbeing | It would be really hard...I would definitely crack after a week and a half or something like that (Aged 31-35, White British) |
|  | Financial implications | So if you're not getting that support financially, and it's not also a rule for everyone to do, then I doubt many people would do it (Aged 21-25, White British) |
|  | Living conditions | I think self-isolation is tricky. If we go back to COVID we can see why it was tricky. One I think based on people's living conditions, you might not live in a space that will allow you to self-isolate (Aged 26-30, White British) |
| Avoidance of activities considered high risk (e.g. attendance at saunas or sex-on-premises venues) | Impact of COVID-19 | If this had happened in 2019 I think it would be so different. I think people would engage, people are just so fed up (Aged 36-40, White British) |
|  | Importance of maintaining social and sexual connections | Just carrying on as normal. I can't hide myself away because there's monkeypox, you can't really hide away. COVID is still here, and it's going to be here, we can't lock ourselves indoors all day, we've got to go out and just live normally (Aged 56-60, White British) |
|  | Low perceived severity | I will actually just put my head in the sand and count on the fact that for a lot of people it hasn't been that severe, maybe it's just going to be like another STI, we take risks all the time with whatever, herpes, syphilis, you know, how different is it? (Aged 31-35, White British) |
| Vaccination | Availability | I've been trying to get the Monkeypox vaccine for a long time now and they still keep saying there's a shortage there's more coming in early September and it's already early September and I'm still waiting for the call. I've tried calling the NHS, I've tried calling [sexual health clinic] and it's just very hard to acquire the vaccine (Aged 36-40, White British) |
|  | Resources and time constraints | There's all these pop-up clinics and because I'm working as well I'm not just able to take time off to go to these pop up clinics and most of these pop up clinics are not around where I work so I even have to take an absence of leave for one day and just go but wouldn't look good in the company I'm working for (Aged 36-40, White British) |

### Sexual health-related knowledge, practices and support

Some interviewees described already being proactive in maintaining sexual health, including regular STI testing and the enactment of seroadaptive behaviours (e.g., partner selection/strategic positioning based on HIV status) to reduce STI/HIV risk to themselves and others. Measures and risk-reducing behaviours in response to Mpox were therefore considered highly acceptable and effective as they aligned with established and ‘normalised’ sexual healthcare routines (*As a gay man who has regular check-ups, it becomes an everyday part of your life…it’s just one of those things that you do to stay safe*, Aged 41-45, White British). High levels of health literacy and engagement with sexual health services also enhanced comfort in one’s ability to detect Mpox and seek appropriate help if required:

> *It would work for me because I also check myself, I’m very aware of my body whether it’s suffering or catching something, so I will easily make a call to my GP or seek advice or help if I catch it - I know who to call if something happens to me* (Aged 36-40, White British)

Those who described barriers to accessing sexual health services or did not report receiving information via social media and GBMSM networks reported how some measures, particularly symptom recognition, were less straightforward to perform. This was because these channels were key sites for public health agencies, community organisations and GBMSM to communicate Mpox health promotion information as well as norms around sexual health and protection. Those who were inadequately represented in and faced difficulties accessing these networks, including racially minoritized groups or people whose first language was not English, therefore had limited exposure to health promotion information, including those communicating Mpox symptoms:

> *…networks that exist on social media are very biased and they are leaving behind those who don’t speak English, who don’t have access to the internet or do not have access to those networks, and that tends to be people who need most support, because they’re the most vulnerable…[we are] generating a mass of white, highly educated people who will rapidly identify their symptoms and are we leaving behind a vast amount of people who don’t know that much about Monkeypox and will not identify it and don’t know they have it* (Aged 26-30, White Other)

Symptom recognition was therefore challenging in the absence of accessible and culturally appropriate information. A particular issue was distinguishing Mpox symptoms from other STIs or common sexual health complaints or injuries. Symptoms that did not match with official guidance emphasizing only ‘severe’ and specific Mpox symptoms (e.g., lesions), undermined the ability to recognise and seek help for potential infection:

> *So I just started sweating, high temperatures, and then I remember about four days into me feeling ill, I started bleeding from my rectum…I thought it’s probably the guy that I had sex with, [because] back then…bleeding from the rectum wasn’t really a symptom* (Aged 26-30, Black Caribbean)

### Stigma and sexual orientation openness

Generally, measures requiring disclosure (e.g., testing, contact tracing) were less acceptable to those either subject to or concerned about Mpox-related stigma. These conversations related to perceptions that they would be seen as ‘dirty’ due to the visual symptoms of Mpox *([I] couldn’t help but feel dirty because it affects your skin, it shows up on you…you couldn’t help but feel stigmatised,* Aged 36-40, White Other), or as someone engaged in ‘irresponsible’ or ‘promiscuous’ behaviours *(all my friends that had it…we all kept it hush-hush from each other, ‘cause it’s sort of like you don’t want to be seen as being the slut,* Aged 26-30, Black Caribbean). These issues were most pronounced among racially minoritized groups or those from geographical spaces with small GBMSM scenes, who were concerned about being identified as a source of transmission in their communities:

> *But also nobody wants to – especially a small city like mine - to be known as one of the first people to have caught it and everybody else thinking that they’re the one that’s the super spreader or whatever. Again, that goes back to the stigma side of things* (Aged 26-30, White Other)

These concerns limited engagement with measures, ranging from the complete avoidance of those requiring full disclosure (e.g., healthcare seeking) (*I already had my own little scare, a couple of weeks back and I was like, ‘Have I?’ I didn’t get tested because I was riddled with shame about it,* Aged 26-30, White Other) to partial engagement or adherence with measures that could be adapted or modified to avoid disclosure (e.g., 21-day isolation). For example, to avoid ‘awkward’ conversations with employers or others who may question *why* they were isolating for a prolonged period, some described isolating only when symptoms were visible or noticeable to others. One interviewee described how they returned to work shortly after symptoms had disappeared, but enacted precautionary measures to limit transmission to others:

> *It was selfish – well, it is awkward – but I went back to work…which I probably shouldn’t of but I did but.. It wasn’t my face – it was locked up in underwear, so I knew that I wasn’t gonna spread it! I just put on a facemask – just to be on the safe side* (Aged 26-30, Black Caribbean)

Those who were open about their sexuality described these situations and conversations as being more straightforward to perform. These individuals were actively involved with LGBTQ+ networks and comfortable about informing others about their sexual practices and behaviour. In contrast to the account above, one interviewee described how his openness removed any potential discomfort about informing his employee why they were isolating:

> *I’m a loud, proud, outspoken gay person who doesn’t feel any kind of shame about having a lifestyle that involves loads of shame. So I was telling work I can’t come in because I could have monkeypox. I’ve got a job that is an office job that are now wanting us to come into the office so I go whole weeks without coming in* (Aged 26-30, White British)

### Retaining Intimacy

A small number of interviewees – particularly those whose attendance at sex-on-premises venues and spaces was central to their social lives and sexual identity - outlined the importance of maintaining social and sexual interactions during this period. For these, the potential cost of a positive test (e.g., 21-day isolation) limited engagement with healthcare services if symptomatic. These decisions were influenced by previous COVID-19 experiences, including how social distancing and the closure of gay venues had restricted opportunities to connect with others through these spaces. In attempts to actively avoid similar experiences, several interviewees described enacting behaviours that allowed for the continuation of social and sexual practices recognised as ‘high-risk’. These ranged from fairly straightforward risk-reduction practices, including limiting sex to known contacts (*I’m not fully getting in touch with new people on the apps, I’m only sticking with people I already know,* Aged 31-35, White Other), to informed-decision making about what sexual practices, and with whom, were permissible under heightened conditions of risk:

> *We’d been in situations in [city] where we’d had a lot of fun, we’d attended a couple of parties, we went to huge club events where there was like 30,000 shirtless men in close proximity. We went to a couple of sex on premises events and stuff like that. So, we knew we’d been at risk but we hadn’t been reckless either. You know, we were quite aware that this was a thing. We were kind of, to the best way that we could, we were checking people* (Aged 36-40, White Other)

### Optimisation of Messaging

Interviews provided in-depth insights into perceptions of UKHSA messages and communications and ways these could be optimised to overcome some of the aforementioned behavioural barriers. All feedback was collated in a table of changes (Table 3) and any potential optimisations and changes to content were discussed during interviews. Although a range of suggestions to the content were made, we have focused on higher priority feedback considered most important in prompting engagement with content and protective behaviours.

**Table 3.** Summary of participant feedback concerning UKHSA messaging and guidance

| Theme identified | Example evidence | Suggested changes based on feedback |
| --- | --- | --- |
| Focus on severe symptoms in messaging could mean that early infection goes unnoticed | People probably think, 'I'm looking for these pimples or these spots or these boils on my hands,' rather than thinking about the milder symptoms. I think that is potentially dangerous because it may not – people might not catch it sooner and continue to put themselves and others at risk. Then we're going to get a spiral of infections (Aged 26-30, White Other) | Provide examples of all symptoms. Suggest in messaging that 'waiting and seeing' or second-guessing symptoms is risky (e.g., "get checked immediately if you have symptoms (even if mild), don't leave until it's too late"). |
| Lack of information regarding Mpox symptoms, including potential symptom overlaps and information on how to differentiate from other STIs | But it also was during the time when it was really, really hot. Loads of insects and bugs everywhere, so I just thought it's definitely that, rather than actually giving myself a scare to go and get tested. Also, I didn't have any of the other symptoms like headache, nausea, anything, (Aged 26-30, White Other)<br>I don't fully know the timeline of when you're meant to see symptoms, or even if these all hit at once, I don't really know to be sure (Aged 26-30, White British) | Provide simple messages emphasizing not only what Mpox symptoms to look out for, but also highlighting potential overlaps with other STIs/infections. |
| Lack of targeted messaging for those at increased risk | I think like with a lot of things, the attitudes kind of evolve. So at the beginning when it was just kind of four cases or five cases and there was a bit of speculation then, obviously about who it was affecting. I think in one way the approach has been right in the sense that at the beginning the messaging was very much about anyone can get it but then, as we know today, it is you know 99% in kind of more of the gay population. So I think myself and most of the people I know are comfortable with that and happy with that kind of focus (Aged 21-25, White British) | Use targeted messaging in specific locations/venues (e.g. GBMSM venues) to reach at-risk populations.<br>Justification for focusing on at-risk should be supported with evidence (e.g., "although Mpox can affect anyone, most cases are among GBMSM").<br>Be mindful of needs of groups subject to pre-existing stigma who may be less amenable to targeted approaches. |
| Absence of harm reduction messages | they won't listen to messaging necessarily, especially if you go full-on don't have sex, I don't think that message is helpful (Aged 36-40, White British)<br>What I think would make so much sense for the Public Health messaging to include is 'this is probably not a good idea to be doing that right now. Have as much | Abstinence-based messages unlikely to have impact (e.g. "do not have sex"). Instead, use harm reduction messages to help people understand risk and reduce potential transmission risks. |
|  | *casual sex as you want but do it in a way that you get contact details.' So if you meet someone through Grindr, save them in Grindr or get their phone number (Aged 26-30, White British)* |  |
| Official posters/messages not as credible as those from within GBMSM community | *And I think probably...this messaging probably wouldn't be able to come from anyone else outside the community. But I think if people within the community were kind of saying that this is looking after the community and like we're in it together in a sense then people would feel more likely to oblige I think. Instead of more top down government saying that gay people have to isolate which naturally I think would make that a bit trickier (Aged 21-25, White British)* | Use experts within the community to provide convincing and credible messages to reach diverse groups at risk populations.<br>Co-produce messages with community-based organisations.<br>Use community-based groups and organisations to disseminate official messages among at-risk communities. |
| Absence of information around looking after yourself with Mpox | *What is the advice to people to look after themselves? Whilst we're at the very beginning of what could be a very significant wave of something which also can have very significant impact on people's lives (Aged 26-30, White Other)* | Include information on self-care, including measures that can be enacted to prevent or lessen Mpox symptoms. |

### Targeted vs Inclusive Messaging

Discussions regarding the delivery and framing of Mpox messaging focused heavily on whether this should be targeted towards those most at risk of contracting Mpox. Some felt that tailored messaging specifying that sexually-active GBMSM were primarily at risk of, and responsible for (through sexual contact), Mpox could be leveraged to further reinforce homophobic tropes and stereotypes of sexual promiscuity and recklessness. In particular, concerns were raised about emphasizing the role of gay sex in transmission, particularly as group sex and anonymous hook ups were already highly stigmatised gay sexual behaviours. Interviewees who were less open about their sexuality, or reported concerns about being judged by others, therefore favoured messaging that avoided linking Mpox acquisition with gay sex (e.g. “Anyone can get it”). These messages limited the potential for further shaming and marginalization:

> *I don’t think it [targeted messaging] would necessarily be more helpful because then it’s causing some kind of stigmatisation or discrimination so it can do more harm than good. So even though the percentage is 90% of cases, but really anyone can still get it so that really is the message* (Aged 26-30, Black Caribbean)

These views contrasted with those actively engaged and participating in online and offline GBMSM networks, who were already proactive in sharing Mpox information and support across these spaces. For this group, generic and non-specific messaging did not accurately reflect the fact that sexual encounters between gay men were the dominant mode of Mpox transmission. Messaging failing to acknowledge the role of sex between men in transmission was therefore inaccurate and risked failing to reach those groups most at risk of contracting Mpox. One interviewee reported how, in the absence of such messaging, targeted peer-to-peer information communicating the role of sex between men was welcomed and helped influence risk perceptions:

> *I don’t think they should sway away from making sure and making it prominent in their reporting that it is spreading amongst men who have sex with men, because that’s the reason that I’m so aware of it now… And I don’t think it’s wrong to report it that way, because it has raised so much awareness between me my gay friends* (Aged 21-25, White British)

### Content and Information

There was a consensus that messaging needed greater clarity and precision in communicating specific and up-to-date Mpox symptoms. This was especially important during the initial stages of the outbreak when information regarding transmission risks and symptoms was often inconsistent and incomplete. As a result, some interviewees reported difficulties distinguishing Mpox symptoms from common STIs with symptom crossovers during these periods (*Well, I don’t know because If it was a rash, I actually have psoriasis as well, so if I thought it might be psoriasis I’d probably ignore it, to be honest,* Aged 21-25, White British). Others felt that messages focusing on ‘severe’ symptoms meant minor ones would go unnoticed. Acknowledging the changing – and atypical – context was cited as a useful method to improve symptom awareness and recognition:

> *I think what they need to do really is to make it clear that this outbreak is atypical, that this outbreak is not what they have seen in Africa…that the one here in the UK is different, it’s more localised in the body and people might not get all the symptoms, they might not even get a rash…so, just to make it obvious that even though it’s the same virus it behaves differently here for some reason* (Aged 41-45, Latin American)

Amongst all interviewees, there was consensus that harm reduction measures, including information on how to reduce one’s risk of acquiring Mpox, would be useful in messages. Abstinence-based measures were perceived as ineffective and unrealistic to perform, and there was a preference for messages promoting the uptake of ‘safer’ behaviours:

> *The idea of telling people to not have sex just doesn’t make any sense because people are not going to in a sense follow that. I think we have to come up with if you’re not made to stop having sex how can we in a sense reduce your risk or make it safer for you to have sex…the message may be reducing your sexual partners* (Aged 26-30, Black Caribbean)

### Communication channels and pathways

An important finding in itself was that most interviewees had not seen UKHSA posters/messages before participating in the study. Instead, most Mpox information had been obtained through social media platforms (specifically Tik Tok and Twitter) or peer-to-peer communication, including information about Mpox symptoms and prevention measures (e.g. vaccination sites). Information obtained from key organisations and voices within the LGBTQ+ community, including the sharing of experiences of Mpox infection and self-care, were perceived as relatable and helped communicate the possibility - and risk - of Mpox infection:

> *I follow some people on Twitter and they posted images of their sores and lesions and it’s quite horrific to say the least, and being gay myself there’s this worry that I will get it in the future as well* (Aged 36-40, White British)

However, there was a perception that communication via social media had a tendency to relay personal and subjective (and often extreme) Mpox experiences that increased the possibility of misinformation being spread. In these situations, official advice from ‘credible’ sources was considered helpful to mitigate information inconsistencies, even if it was incomplete or lacking in specific detail at the time:

> *I think that sometimes you need an official source, you look to people like UKHSA as kind of the bastions of the facts and I think that when people like that are not talking, when NHS England are not telling you anything about monkeypox, when the media are only reporting bad things about it and making out like it’s some new gay plague…[they] are super important because they go, ‘Right, here’s the facts. This is what we know. This is all we know right now and it’s probably not everything that you want to know but we are telling you everything that we know.’ So, it’s an honest, truthful version* (Aged 46-50, White British)

## Discussion

Our study offers key insights into factors influencing the intended and actual uptake of protective behaviours among GBMSM during the 2022-23 Mpox outbreak in the UK. First, we identified the importance of Mpox risk perceptions, finding that those engaging in high-risk sexual practices were motivated to engage or enact some mitigation measures, despite low perceptions of Mpox severity. Second, mitigation measures were perceived as acceptable if they fitted within pre-existing risk-reduction practices. Given how connections to GBMSM networks and social media channels were found to increase exposure to sexual health information and norms influencing protective behaviours, those excluded or marginalized from these networks were less aware of sexual health promotion behaviours and specific Mpox mitigation measures (e.g., symptom recognition). Finally, our findings examine how public health communications can be optimised to improve the uptake of public health measures, particularly among GBMSM excluded or marginalised from the key sites of information identified in our study, and sub-populations of GBMSM engaged in sexual practices and behaviours that increase transmission risks.

We found that both intended and actual engagement with public health measures were influenced by prior engagement in activities considered to be high-risk, including group and frequent casual anonymous sex. Those managing risk through monogamous sexual relationships perceived certain mitigation measures (e.g., vaccination, healthcare seeking) as therefore unnecessary or redundant. These findings support quantitative data detailing how heightened perceptions of personal risk and proximity to a viral threat, including Mpox, are associated with willingness to adhere to mitigation measures (18, 37). Whilst this demonstrates that most GBMSM were well informed of Mpox transmission mechanisms, younger interviewees and PrEP users reported being less concerned about the severity of Mpox, corresponding with findings showing lower Mpox severity perceptions among PrEP users compared to non-PrEP users (38). Although this could be a consequence of vaccine efforts in the UK initially focusing on this group (thereby resulting in higher perceived protection), decreased perceived susceptibility to HIV/AIDS due to PrEP use may outweigh concerns of acquiring ‘curable’ bacterial STIs (39–41), a finding expressed by some PrEP users in our study in relation to Mpox. These findings are disconcerting given recent reports of severe Mpox infection among PLHIV (42) and highlight the need for increased education around the severity of infection on one’s health. Public health communications considering different levels of perceived severity among different groups have been shown to be effective in changing determinants of behaviours during viral outbreaks (43), and similar methods that communicate the severity of Mpox infection to those who consider this to be low may be suitable.

Social media and GBMSM networks and spaces were key domains for disseminating Mpox health promotion materials, including information about symptoms, vaccine availability and risk reducing behaviours. As such, connections to these networks influenced one’s ability and capacity to follow mitigation measures, including those dependent on specific and timely information around symptoms (e.g., healthcare seeking) and those influenced and prompted by social norms and responsibilities (e.g., vaccination, isolation, risk reducing behaviours).

However, in line with studies exploring the uptake of HIV self-testing (44) and PrEP (45) among GBMSM, we found how social and geographical marginalisation from these spaces limited exposure to information and community norms regarding Mpox protective behaviours. If sexual health promotion campaigns seek to utilise these networks for the delivery of public health information^1^, a key issue will be to therefore consider the complex set of intersecting dynamics (e.g., discomfort, racism, stigma, gendered norms) that inhibit some groups connections with these spaces (44). Issues of limited exposure to and experience of enacting behaviours, as observed in our study, suggest that additional support is also required to improve health literacy and self-efficacy in enacting behaviours among those groups (43).

Related, those who felt stigmatised and closeted about their sexual orientation reported difficulties in adhering to measures requiring disclosure, including healthcare seeking, contact tracing and isolation. We also identified how stigma directly related to Mpox, including its visibility and perceived associations with ‘promiscuous’ behaviour, served to inhibit people from adhering fully to guidelines even when risk is known to be high.

Examples from HIV and other STIs highlight similar detrimental effects of stigma on medical adherence, including contact disclosure (46) and testing uptake (47). As in our work, this is particularly acute among GBMSM from minority ethnic (45) and/or non-urban backgrounds (48). In this context, it is helpful to draw upon insights and lessons learnt from HIV/AIDS and COVID-19 to help inform stigma-informed pandemic responses that seek to address stigma experiences and healthcare access among affected communities (49). This includes practical measures (e.g., financial support and reimbursement (50)) supporting the uptake of isolation, as well as health promotion activities and messages combating harmful narratives (e.g., role models and ambassadors directly calling out stigma-inducing information (51)). As with HIV responses, any future pandemic preparedness and response measures to reducing stigma should be co-designed with affected communities to ensure they target heterogeneous experiences and drivers of stigma among different populations of GBMSM (52).

Although the majority of our interviewees reported reductions in sexual activity during this period, we identified a small sub-section of interviewees continuing to engage in sexual activity and practices recognised as high-risk. These findings are consistent with those from COVID-19 suggesting some GBMSM may find it challenging to fully adhere to social distancing and isolation guidelines if they retain a desire for physical and sexual intimacy during quarantine/lockdown periods (53, 54). Although these interviewees were not following public health guidance, our findings align with a body of research documenting how GBMSM engaged in ‘high-risk’ sexual behaviours independently and carefully deploy risk-reduction techniques in efforts to negotiate viral threats (53). For example, we identified similar practices whereby some interviewees reported limiting sex to known contacts or making careful decisions about what social and sexual behaviours, and with whom, were permissible during this period. Given context-driven and personable health information in public health messaging has been shown to be more effective in changing behaviours (43), learning from and engaging with these practices and techniques will be beneficial in developing interventions and prevention measures that meet the needs of sub-populations of GBMSM formed around particular social and sexual behaviours (53).

Finally, our findings also uncovered important preferences for communicating and disseminating information during the outbreak. For some interviewees, the inclusive approach to messaging taken by UKHSA was inconsistent with the reality of Mpox disproportionally affecting GBMSM. Preferences for socially targeted messaging that highlighted the role of sex between men in transmission were therefore considered accurate, evidence-based and capable of reaching groups most at risk of Mpox acquisition who may fail to recognise themselves in non-specific messages. Although these findings suggest targeted messaging may be effective in maximising relatability (55), concerns were also expressed about the potential of targeted approaches in reinforcing stigmatising and negative depictions of gay sexual practices. These findings accord with mixed public responses to a targeted and sex-positive messaging campaign aimed at increasing PrEP awareness among sexual, gender and racial minorities (*PrEP4LOVE*) (56): despite increased community knowledge and awareness of PrEP, targeted promotional material was found to exacerbate converging forms of stigma related to ethnicity and sexual orientation. Given preferences for inclusive messaging in our study were predominantly among either GBMSM of minoritized ethnic backgrounds or GBMSM concerned about judgement related to their sexual orientation and behaviours, these findings suggest GBMSM may have particular preferences for messaging based on their previous experiences of stigma and discrimination. Messages should therefore be co-created alongside specific populations of GBMSM to reflect heterogeneity in experiences.

The majority of interviewees were in favour of the UKHSA approach that relied upon community engagement in risk communication, although the aforementioned finding that those excluded or with weaker network ties were less exposed to information suggests risk communications may need to move beyond these spaces to enhance awareness, education and knowledge of Mpox among these groups. Further, whilst communications through these networks ensured relatability and heightened perceptions of personal risk (43) there was the potential for ‘uncontrolled’ sources to communicate misleading or exaggerated information (e.g., a focus on severe cases). In these instances, timely and honest information from official sources that acknowledges inaccurate information can improve public trust and acceptance of recommended behaviours (43, 57). This option is supported by our findings, as many interviewees indicated that they would like to receive more honest and timely information from official sources to dispel concerns about exaggerated or misleading information.

Our study has a number of strengths and limitations. The use of a rapid qualitative methodology enabled us to quickly explore actual rather than self-reported intended behavioural responses, which do not always translate into enacted behaviours (50). The qualitative interviews also provided in-depth understandings of GBMSMs’ social contexts and how these influenced adherence to mitigation measures. Combining this with a co-production element enabled us to identify potential optimisations that can maximise acceptability of interventions and messages. These may be of interest more widely to those developing and maximising the acceptability of public and sexual health interventions for GBMSM in response to new and emerging viral threats. Our recruitment methods, that relied upon key community organisations for distribution in both online and offline LGBTQ+ spaces, also enabled us to reach populations of GBMSM at most increased risk of Mpox, including those who found Mpox advice difficult to understanding or those engaging in sexual practices considered high-risk. Our study also included a sizeable portion of interviewees who were from minoritized ethnic groups or whose first language was not English, which allowed for comparisons and understandings of the differential Mpox experiences among groups who face additional barriers to healthcare services.

Nevertheless, despite targeted recruitment efforts the study is limited by the over-representation of GBMSM with higher educational qualifications, healthcare involvement and health literacy. Studies have shown that those with similar characteristics have higher vaccination rates against Mpox (18), suggesting these groups are more likely to engage with protective behaviours. Similarly, whilst our study includes a sample made up of interviewees from minoritized ethnic groups, only a small number of interviewees represented each ethnic group involved in this study. We were therefore unable to identify and make strong inferences regarding different levels of Mpox understanding and healthcare access both within and across ethnic groups. Our findings are also predominantly representative of the views of cis-male interviewees, and there is a need to understand the views of both nonbinary and trans persons who have sex with men (TPSM) whose experiences may also differ.

Finally, interviews were conducted over several months and views and experiences of Mpox may have changed as new information regarding symptoms and severity became available. For example, recent evidence of severe Mpox infection (42) may have altered the low severity perceptions evidenced in our findings. The timing of interviews therefore require consideration when interpreting the findings.

## Conclusion

Findings highlight the importance of risk perceptions in shaping both intended and actual behaviours toward public health measures during the 2022-23 Mpox outbreak. Barriers related to low health sexual health literacy (and exclusion and marginalization from key sites and spaces for obtaining information), stigma and the importance of maintaining sexual and physical intimacy limited adherence and acceptability among some groups of GBMSM. To help ensure equitable access among different groups of GBMSM, future sexual and public health measures deployed in response to emergencies will need to be attentive to how the differential needs, preferences and experiences of GBMSM limit the acceptability of measures. Key to this will be the co-design of public health interventions and campaigns in consultation with diverse groups and communities, to ensure greater acceptability and credibility in different contexts and communities.

## Data Availability

The dataset is qualitative and includes numerous highly sensitive quotes from which participants are potentially identifiable. For this reason, the raw dataset will not be available. Additional quotes supporting each theme will be made available on request.

## Acknowledgments

TM, SD, JH, MH, RA, IO and LY are funded by the NIHR Health Protection Research Unit (HPRU) in Behavioural Science and Evaluation at the University of Bristol, a partnership between UKHSA and the University of Bristol.

LS, GJR and RA are funded by the NIHR Health Protection Research Unit (HPRU) in Emergency Preparedness and Response at King’s College London, a partnership between UKHSA and Kings College London.

LY is an NIHR Senior Investigator and her research programme is partly supported by NIHR Applied Research Collaboration (ARC)-West, NIHR Health Protection Research Unit (HPRU) for Behavioural Science and Evaluation, and the NIHR Southampton Biomedical Research Centre (BRC).

JH is also supported by NIHR Applied Research Collaboration (ARC)-West

The views expressed are those of the authors and not necessarily those of the NIHR, UKHSA, or the Department of Health and Social Care.

We would like to thank the Terrence Higgins Trust and PrEPster for their support with participant recruitment.

## Contributions

Conceived the study: TM, LS, JH, SD, GJR, RA, IO, LY. Study design: TM, LS, JH, SD, GJR, RA, IO, LY. Analysed the data: TM, LT. Interpreted the data: TM, LT, LS, JH, SD, GJR, MH, RA, IO, LY. Drafted the manuscript: TM. Reviewed the manuscript and approved content: TM, LT, LS, JH, SD, GJR, MH, RA, IO, LY. Met authorship criteria: TM, LT, LS, JH, SD, GJR, MH, RA, IO, LY. The author(s) read and approved the final manuscript.

## Ethics decelerations

### Ethics approval and consent to participate

Ethical approval for this study was granted by UK Health Security Agency Research Ethics and Governance Group (ref R&D 512). All participants provided informed consent/assent to participate. All methods were carried out in accordance with relevant guidelines and regulations.

### Consent for publication

N/A

## Footnotes

1 The UKHSA strategy for disseminating messaging relating to Mpox largely relied upon a network of LGBTQ+ and sexual health organisations (including third-sector and voluntary organaisations) to support engagement among at-risk and marginalized groups.

*** END ***

