## Supplementary material for "Mpox knowledge, behaviours and barriers to public health measures among gay, bisexual and other men who have sex with men in the UK: A qualitative study to inform public health guidance and messaging": Supplementary material.pdf

**Indicative draft interview schedule for Monkeypox co-production**

- Intro to project
- Any questions?
- Recap of information sheet/consent form
- Explain what will happen (we will ask for views of monkeypox in general, then introduce guidance, ask for opinions, both positive and negative, and views on what might be helpful to improve guidance)
- Explain that they do not have to answer any questions
- Check happy to record

vaccina
*Questions*

- Can you tell me what you know/have heard about Monkeypox?
- How serious do you think monkeypox is for you? For those around you?
- **How concerned are you about catching monkeypox? About those around you catching monkey pox?**
- Can you tell me anything you know about current health advice about reducing the spread of Monkeypox?

*Introduce each specific behavioural element of UKHSA guidance and repeat questions for each of:*

- *healthcare seeking/testing*
- *disclosing contacts*
- *self-isolating*
- *avoiding/isolating pets.*
- *getting vaccinated.*

***Where existing materials are available then suitable length excerpts can be used as stimulus materials to present guidance (e.g. advice on what to do if suspect monkeypox).***
*If asking about guidance element in principle (e.g. if relevant guidance not yet issued) then for each target behaviour describe the required behaviour as precisely as possible (e.g. testing immediately notice any symptom; disclosing all contacts; self-isolating for full 21 days with no contact with anyone).*

- What do you think about this advice?
- How would you feel about trying to follow the advice? *(adapt all items as necessary for confirmed cases, e.g. 'How do you feel')*
- What do you think would make it difficult to follow the advice? *(has there been a time when you haven't followed the guidance?)*
- What do you think would make it easier to follow the advice?
- What do you think people you know would think of the advice

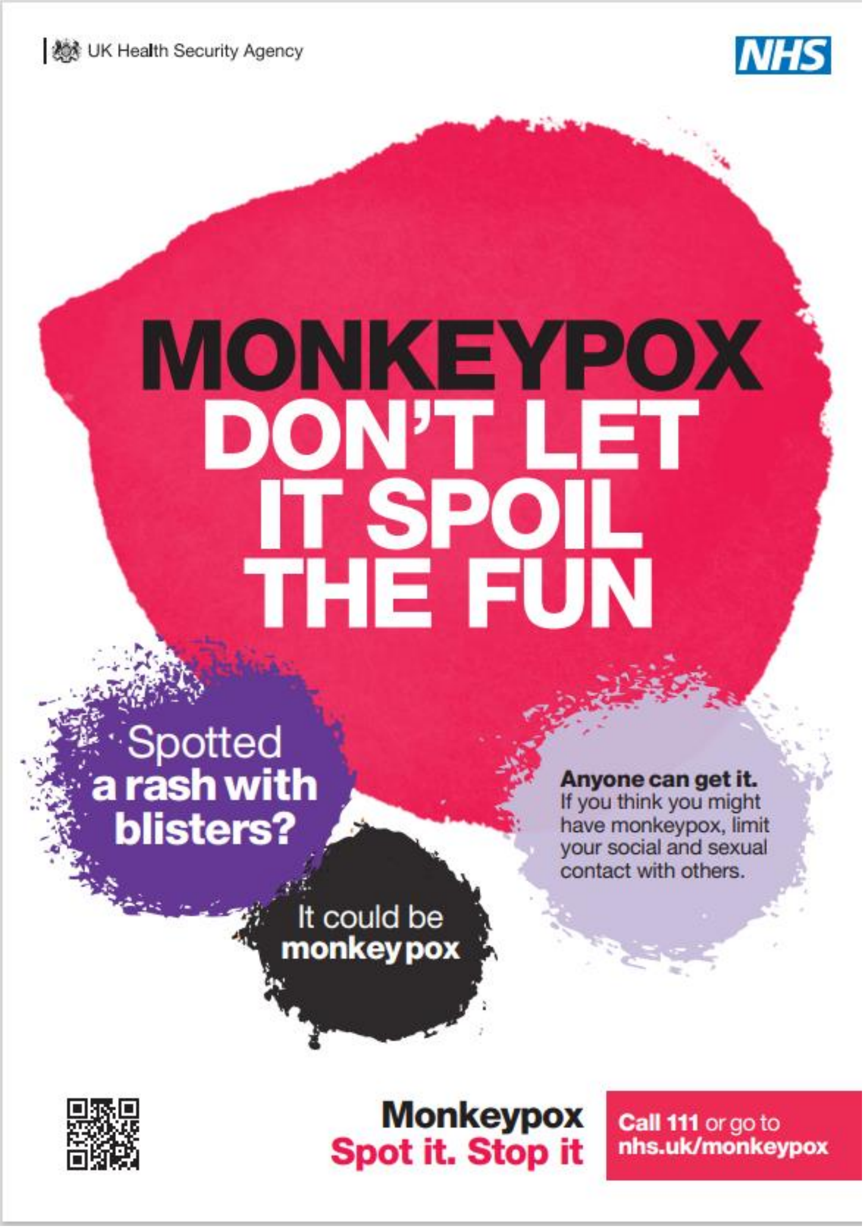
UK Health Security Agency
NHS
MONKEYPOX
DON'T LET IT SPOIL THE FUN
Spotted a rash with blisters?
It could be monkeypox
Anyone can get it.
If you think you might have monkeypox, limit your social and sexual contact with others.
Monkeypox
Spot it. Stop it
Call 111 or go to nhs.uk/monkeypox

UK Health Security Agency

NHS

### MONKEYPOX DON'T LET IT SPOIL THE SUMMER

**What is Monkeypox?**

**Anyone can catch it.**

Monkeypox is a viral infection that spreads through close skin-to-skin contact (including kissing and sex), and by sharing items like bedding and towels. And while it's not life-threatening, it can spoil your fun by making you unwell - and infectious to other people - for several weeks.

**What are the symptoms?**

- A nasty rash with blisters, spots or ulcers that can appear anywhere on your body (including your genitals)
- Fever
- Headaches and muscle aches
- Swollen glands
- Chills and exhaustion

**How can you stop it from spreading?**

If you, or anyone you've been in close contact with, see an unexpected rash with blisters, spots or ulcers anywhere on your body:

- Call 111 or go to nhs.uk/monkeypox as soon as you can
- Avoid close physical contact with others until you've had medical advice

**Monkeypox**
**Spot it. Stop it**

**Call 111** or go to **nhs.uk/monkeypox**
